# A Protocol to Implement and Evaluate a Community Walking Blood Bank for Hemorrhagic Shock When Banked Blood is Unavailable - The Local Initiative For Emergency Blood (LIFE-Blood) Study, Phase 2

**DOI:** 10.1101/2025.05.27.25328015

**Authors:** Nikathan Kumar, Fleming Mathew, Riya Sawhney, Avery Thompson, Cindy Makanga, Wendy Williams, Caroline Wesonga, Abdirahman Musa, Hillary Barmasai, Maurice Wakwabubi, Elizabeth Wangia, Cavin Opiyo, Epem Esekon, Gilchrist Lockhoel, Juan Carlos Puyana, Pratap Kumar, Meghan Delaney, Nobhojit Roy, Andrew P. Cap, Tanujit Dey, Stephen Onyango, Edward Mutebi, Nicodemus O.M. Nyanamba, Kristina Kuya Lokuruka, Kevin Hongo, Nakul Raykar, Tecla Chelagat, Linda Barnes

**Affiliations:** Program in Global Surgery and Social Change, Department of Global Health and Social Medicine, Harvard Medical School, Boston, MA, USA UCSF East Bay General Surgery, Oakland, CA, USA; Division of Thoracic Surgery and Interventional Pulmonology, Beth Israel Deaconess Medical Center, Harvard Medical School, 185 Pilgrim Road, W/DC 201, Boston, MA, USA; Program in Global Surgery and Social Change, Department of Global Health and Social Medicine, Harvard Medical School, Boston, MA, USA; Institute of Healthcare Management, Strathmore University Business School, Nairobi, Kenya; Center for Surgery and Public Health, Brigham and Women’s Hospital, Boston, MA, USA; Blood DESERT Coalition, Boston, MA, USA; Department of Health and Sanitation, Turkana County Government, Lodwar, Kenya; Hemovigilance Officer, Ministry of Health, Nairobi, Kenya; KTTA Ministry of Health, Nairobi, Kenya University of Nairobi, Nairobi, Kenya; Senior Health Specialist, World Bank Group, Nairobi, Kenya; Strathmore University Business School, Nairobi, Kenya; Turkana County Government, Lodwar, Kenya; County Chief Officer, Department of Livestock Development, Turkana County Government, Lodwar, Kenya; O’Brien Chair of Global Surgery, Royal College of Surgeons in Ireland, Dublin, Ireland Professor of Surgery, University of Pittsburgh, Pittsburgh, PA, USA; Institute of Healthcare Management, Strathmore University Business School, Nairobi, Kenya Global Business School for Health, UCL, London, England; Children’s National Hospital, Washington DC, USA George Washington School of Medicine & Health Sciences, Washington DC, USA; WHO Collaborating Centre for Emergency, Critical and Operative Care, The George Institute of Global Health, New Delhi, India University of Global Health Equity, Kigali, Rwanda; Department of Medicine, Uniformed Services University, Bethesda, MD, USA; Lodwar County Referral Hospital, Lodwar, Kenya; Ministry of Health, Turkana County Government, Lodwar, Kenya; Lorugum Subcounty Hospital, Turkana County, Lorugum, Kenya; Department of Surgery, Brigham and Women’s Hospital, Boston, MA USA Program in Global Surgery and Social Change, Harvard Medical School, Boston, MA, USA; Strathmore University, Nairobi, Kenya; Linda S Barnes Consulting, Seattle, WA, USA

**Keywords:** Community walking blood bank, Emergency transfusion protocols, Effectiveness-implementation study, RE-AIM framework

## Abstract

**Introduction:** Trauma, obstetric hemorrhage, and severe anemia lead to millions of deaths every year. The majority of these deaths occur in regions known as “blood deserts” where there is virtually no access to blood transfusions. Currently, these blood deserts have minimal resources to improve access to this life-saving medicine. A community walking blood bank (CWBB) is a low-resource strategy that can provide just-in-time, point-of-care tested, blood transfusions in blood deserts when banked blood is not readily available and the alternative is almost-certain death. This protocol is designed to evaluate the effectiveness and implementation of a CWBB at Lodwar County Referral Hospital (LCRH) located in a blood desert in rural northwest Kenya.

**Methods:** We will use a mixed methods approach relying on an implementation science design to evaluate effectiveness, acceptability, applicability, and impact of a CWBB. First, a previously developed emergency transfusion protocol will be validated and finalized by key hospital stakeholders. Effectiveness will be assessed 1) quantitatively, using prospective laboratory-based data collection before and after protocol initiation to measure changes in blood ordering practices and 2) qualitatively, through key informant interviews of hospital staff and the community focusing on clinical blood demand and general understanding and perceptions about blood donation and transfusion. Lastly, we will determine the adaptability and scalability of a CWBB to other low-resource settings with in-depth interviews, and a modified Delphi approach to achieve consensus regarding key components of a CWBB and its transferability to other settings.

**Discussion:** The global shortage of blood products represents one of the most pressing challenges in healthcare delivery. This study represents the first structured implementation and evaluation of a community walking blood bank, addressing an urgent need for innovative solutions to chronic blood shortages in remote settings. We will prospectively quantify blood demand, understand the impact of a walking blood bank on healthcare providers’ practices, and lastly, explore how it can be adapted to a variety of settings. Through this implementation-effectiveness approach, we hope to provide evidence for a sustainable solution to otherwise preventable deaths due to chronic blood shortages in remote settings.

**Trial Registration:** Not applicable

## Introduction

Low- and middle-income countries (LMICs) face an estimated annual shortage of 102-million units of blood in their blood banks.[1] This deficit is particularly severe in sub-Saharan Africa and South Asia, impacting millions of lives.[1] In rural settings, the situation is even more dire. Facilities with banked blood are often hours or days away, in urban centers. We call these regions “blood deserts” - areas where virtually no screened blood is available for immediate use as the basic treatment of life-threatening trauma, postpartum hemorrhage, or anemia.[2]

Turkana County in Kenya illustrates the challenges faced in these blood deserts. It is one of Kenya’s most remote, impoverished, and medically underserved regions. Lodwar County Referral Hospital (LCRH) is the only major site of blood transfusion in the county, serving a population of 1.2 million, but lacks a comprehensive blood testing facility. Blood is collected on site, but samples must be sent for transfusion transmissible infection (TTI) testing at a regional center in Eldoret, over 300 km away. The Eldoret Regional Blood Transfusion Center (RBTC) is one of six regional transfusion centers mandated with TTI screening across the country. Testing donated blood typically takes 3 to 14 days from initial sample dispatch to result. Thus, while type-specific blood may be present in the LCRH refrigerator, it cannot be used until the test results are received from the Eldoret Regional Blood Transfusion Center (RBTC) days or weeks later. During these periods, patients may die from blood loss while there is unscreened blood at the hospital awaiting TTI results. The LCRH blood bank struggles to maintain an adequate blood supply for its patients, but this delay exacerbates their extreme blood shortages.

An innovative strategy to address emergency blood shortages gaining international traction is the community walking blood bank (CWBB). This approach involves mobilizing pre-screened blood donors on-demand, in times of emergency, using point-of-care screening technologies with rapid diagnostic testing (RDT) and just-in-time whole blood transfusion.[3, 4] The WBB model has been successfully employed by the U.S. military in austere battlefield settings, several high-income civilian settings, and has been used informally for decades in LMICs.[5–8] However, in many low resource settings, concerns persist regarding the safety of RDT-based TTI screening. Therefore, the WBB model has not been officially used to address chronic blood shortages in community settings in low-resource blood deserts.[9–11]

Our team previously explored the need, safety, and feasibility of a CWBB at LCRH. Our study revealed that: (1) blood was unavailable more than 30% of the time; (2) blood unavailability forced providers to resort to emergency measures including just-in-time transfusion practices where blood donations were obtained from established donors and healthcare workers and collected units were screened using point-of-care, RDT prior to transfusion; and (3) point-of-care RDTs performed well in detecting TTIs (HIV, hepatitis B and C, and syphilis) with a 99.2% negative predictive value based on data collected in 2024 within this setting.[12]

Given our increased understanding of blood transfusion needs at LCRH and potential strategies to mitigate it, stakeholders identified the need for a standardized emergency transfusion protocol (ETP) using a CWBB for obtaining blood when banked blood is unavailable and the patient has a high risk of dying, and that this process should be integrated into LCRH’s practices. The aim of this study is to formalize the ETP, evaluate its effectiveness and implementability at LCRH, and understand its adaptability and scalability to broader health systems within Turkana County, regionally, and internationally.

## Aims and Objectives

## Aim 1

To formalize a context appropriate CWBB - ETP at LCRH that aids clinician decision making when no screened, banked blood is available in emergency situations requiring transfusion.

## Aim 2

a. To evaluate the effectiveness and implementation of the ETP by quantifying change in met demand and time to blood dispatch before and after ETP initiation.
b. To understand health care providers perceptions regarding blood availability and true demand at LCRH to explore potential blood-related stressors and clinical impressions regarding the instituted ETP and its effect on decision making around emergency blood transfusion.
c. To understand community perceptions regarding blood transfusions and blood donations to identify potential barriers and facilitators of CWBB implementation and potential motivations or deterrents of donating blood and receiving transfusions in emergency situations.

## Aim 3

a. To identify adaptations of, and potential barriers to, a CWBB necessary to address blood insufficiency in a variety of settings, including Turkana hospitals without blood collection or storage capabilities.
b. To identify main components and potential modifications to adapt ETPs to a broader regional and international context outside of Turkana.

## Methods and Analysis

This study aims to formalize, implement, and assess the impact of a community walking blood bank (CWBB) emergency transfusion protocol (ETP) at the sole primary county referral hospital, LCRH, in Turkana County, through an effectiveness-implementation hybrid design.[13] This will be conducted in three aims.

## Emergency Transfusion Protocol (ETP) Design

Based on results from the first phase of our study, we developed an ETP to aid in decision making processes when screened, banked blood is unavailable to assist providers in emergency situations where there is a significant risk of death or disability (**Figure 1**). This protocol is activated in two scenarios. 1) There is no blood available for transfusion; 2) Blood is available, but it has not been screened for TTIs (HIV, hepatitis B, hepatitis C, and syphilis). Otherwise, standard transfusion procedures will be followed. In these two situations, the clinical team and blood banking staff will first attempt to obtain blood from nearby centers. If these options have been exhausted and all parties – patient, or decision making proxy, the blood bank, administration, and clinical teams – agree, RDT-based screening and transfusion will be initiated. If no blood is available, blood banking staff will 1) contact the RBTC to see if test results are available, 2) contact nearby health facilities that may have blood available to share, and 3) if these steps fail to provide blood for the emergency, then begin contacting potential donors through a pre-established donor registry, prioritizing repeat donors that have previously tested negative for TTIs. After obtaining blood (either previously collected or obtained from a contacted donor), it will then be screened and prepared for transfusion. Samples will also be sent to the Eldoret RBTC which uses standard-of-care enzyme immunoassay (EIA) testing per the national and international standards. If results are discordant with RDT, then the hospital will follow up with patients and donors, notifying them of results and initiating post-exposure treatment protocols per LCRH’s existing policies.

**Figure 1:**
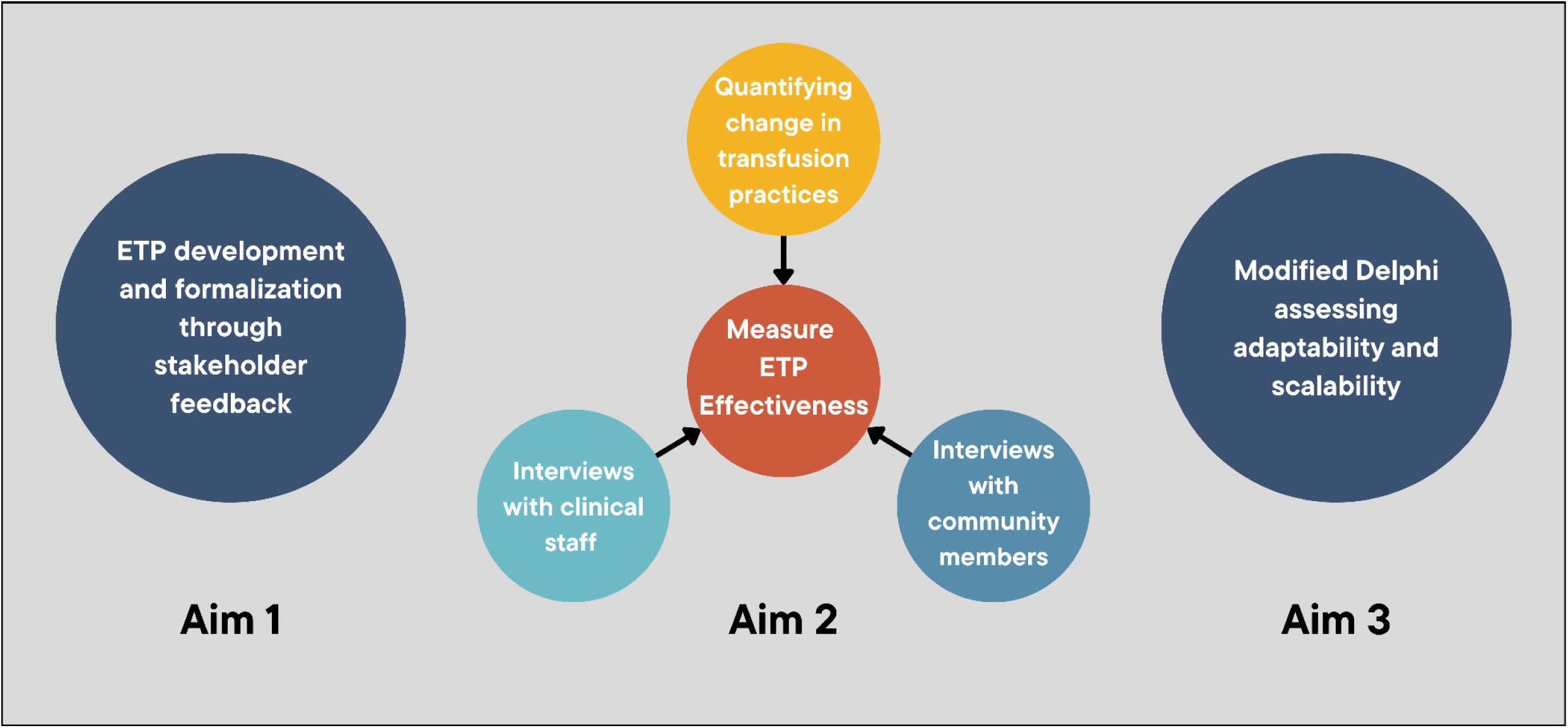
Sequential mixed-methods study to develop a context-appropriate emergency transfusion protocol (ETP) to aid clinician decision-making at Lodwar County Referral Hospital. This protocol consists of three aims: Aim 1 - validate and formalize an emergency transfusion protocol (ETP); Aim 2 - assess the effectiveness of the ETP through quantitative and qualitative research methods; Aim 3 - determine the adaptability and scalability of the ETP through a modified Delphi procedure.

## Study Design

We will begin the study by validating and formalizing the ETP for clinical use at LCRH (Aim 1). Next, we will evaluate the impact of the ETP on transfusion practices at LCRH through both quantitative and qualitative analysis (Aim 2), and finally assess adaptability and scalability within Turkana and more broadly at a national and international scale (Aim 3).

Currently, when a patient is imminently at risk of dying from lack of blood, LCRH mobilizes an unstructured CWBB that often relies on hospital staff, family members, or friends to provide blood in emergency situations using RDTs. To validate and formalize the previously designed ETP process (Aim 1), we will conduct 1:1 stakeholder interviews with hospital and community leadership. We will also engage physicians, nurses, blood bank and laboratory personnel, hospital administrators, and community representatives through larger feedback sessions to build consensus on a finalized ETP. Our objective will be to capture a wide array of perspectives and establish a context-appropriate ETP at LCRH.

Evaluation of the ETP (Aim 2) will use quantitative and qualitative methods to compare two phases of the implementation process: pre- and post-ETP initiation. We will collect data from the laboratory and blood bank to measure met demand for blood transfusion and to understand the timing for key points in blood delivery using the ETP. We will assess blood need and availability at LCRH using short-term, recall-based, cross-sectional surveys of providers at multiple time points pre- and post-ETP implementation. Lastly, we will hold 1:1 qualitative interviews with clinicians and hospital staff to understand their perceptions regarding blood availability as part of the blood transfusion continuum, and with community members, to understand factors behind decisions to donate or receive blood in this applied setting.

This ETP has the potential to be a tool for other low-resource settings with limited blood supplies. Therefore, understanding the adaptability of a CWBB in other settings is critical (Aim 3). To understand the adaptability within the Kenyan context, we will perform key informant interviews and focus group discussions with frontline workers and health administrators in up to three first-level hospitals within Kenya to assess what adaptations may be needed in different settings. We will additionally conduct a modified Delphi process with healthcare leadership at the county and national level within Kenya, as well as an international level with experts from high- and low-income settings to understand multi-level barriers and facilitators of broader CWBB implementation.

## Study Period

The study will be conducted over the course of one year. Qualitative interviews to finalize and validate the ETP (Aim 1) will occur within the first month of the study. Aim 2 will occur over the course of 7 months. Initial prospective data collection will start in tandem with Aim 1 and continue for two months pre-ETP implementation. After two months, the ETP will be initiated and post-implementation data collection will begin, continuing for five months. The short-term recall survey of providers will consist of four, 1-week daily surveys that will occur throughout the prospective data collection phase. One week will occur pre-ETP initiation, while the remaining three weeks will occur separately throughout the post-ETP period. Exploring community and stakeholder perspectives will begin after the ETP has been in place for over four months. Aim 3 will be conducted at the end of the year, with plans to use preliminary data from Aims 1 and 2 to stimulate discussion among key stakeholders.

## Data Collection/Data Sources

Semi-structured interviews will be conducted in each aim of the study to receive ongoing local input and feedback regarding the study and the ETP (**Table 1**). Key informant interviews from Aims 1 and 2 will focus on the context at LCRH and will be conducted with hospital administrators, physicians, nurses, laboratory and blood banking staff, and community members. Aim 3 will draw on the expertise of health officials, doctors, nurses, medical staff, and community members from sub-county hospitals in Turkana where blood is scarce, to focus on local adaptations.

**Table 1:** Interview Guide Matrix. This highlights the representative interview themes and the specific stakeholders that will be involved in each interview. Interviews are delineated by the specific aim.

|  | Hospital<br>administrators | Government<br>representatives | Frontline<br>providers | Laboratory and<br>transfusion staff | Community<br>members |
| --- | --- | --- | --- | --- | --- |
| **Aim 1: ETP validation** |  |  |  |  |  |
| Feasibility | ✓ |  | ✓ | ✓ |  |
| Logistics and flow | ✓ |  | ✓ | ✓ |  |
| Context appropriateness | ✓ |  | ✓ | ✓ |  |
| Implementation pearls | ✓ |  | ✓ | ✓ |  |
| Monitoring | ✓ |  | ✓ | ✓ |  |
| **Aim 2: Provider and**<br>**community perspectives** |  |  |  |  |  |
| ETP integration experience |  |  | ✓ |  | ✓ |
| Impact on patient care |  |  | ✓ |  |  |
| Ethical considerations |  |  | ✓ |  | ✓ |
| Prior blood donation<br>experience |  |  |  |  | ✓ |
| **Aim 3: Essentials for**<br>**expansion and adaptation** |  |  |  |  |  |
| Existing protocols | ✓ |  |  |  |  |
| Impressions of ETP | ✓ | ✓ |  |  | ✓ |
| Context-specific barriers<br>and facilitators | ✓ | ✓ |  |  | ✓ |
| Implementation logistics | ✓ | ✓ |  |  |  |

The research team managing interviews will consist of a senior researcher experienced with qualitative methods, and several research assistants. Interviews will be conducted in English, with Swahili and Turkana interpreters available for interviewees as needed. They will be held in-person, or in rare cases, via online platforms, and expected to last 30-60 minutes. Discussion prompts have been reviewed and finalized by the research team. If further changes are suggested during the interview process, all researchers will have to agree to adopt these changes into the prompts.

A modified Delphi process with a diverse group of health system administrators, clinical providers, ministry of health officials, and non-governmental organization leadership will be conducted to understand national and international adaptations (Aim 3).[14] This will be conducted in at least two rounds to obtain consensus. The validated ETP from Aim 1 will be presented along with a set of questions to elicit feedback response. Feedback will be obtained in real time electronically to ensure anonymity of responses and to avoid group conformity bias.

Quantitative data collection will be limited to Aim 2. Our retrospective data collection will rely on the “crossmatch” and “stockout” registries at LCRH back to six months before the start of prospective data collection. The crossmatch registry is used by the laboratory at LCRH to record all crossmatched blood that has been dispatched to a patient. The stockout registry is a separate listing used at LCRH to denote blood requests that were unmet due to blood stockouts. Prospective data collection will occur within the laboratory at LCRH. We will collect information regarding every blood request over the study period (**Figure 2**). This includes indication, patient demographics, urgency, blood availability at the time of request, and if the ETP is activated, metrics regarding time to dispatch, donor characteristics, RDT results, and concordance with the regional testing site. In addition, we will capture daily quantification of blood supply (by type) in the blood bank. These will be paper forms that will be completed by a designated research member so as not to disrupt clinical activities. We will then digitize the forms for analysis.

**Figure 2:**
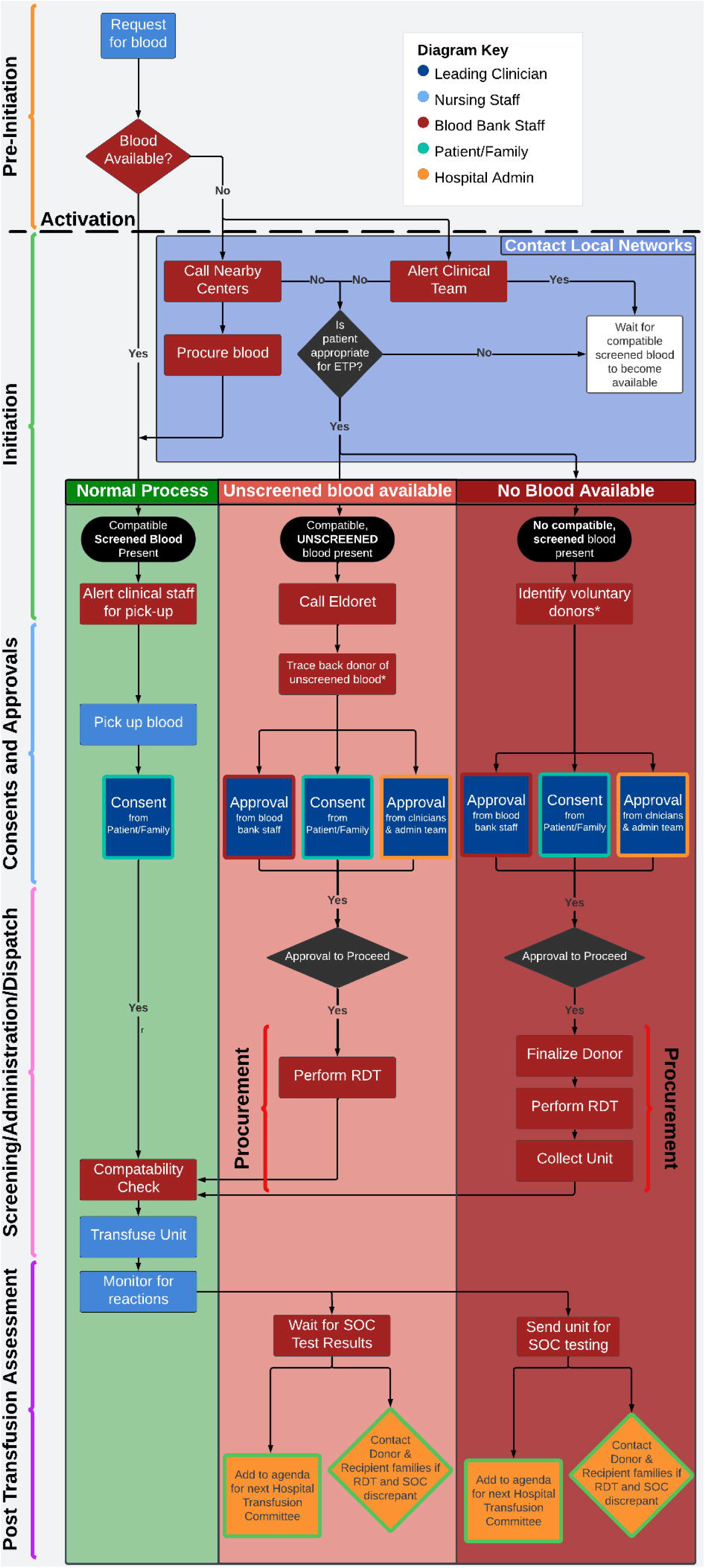
General Flow Diagram and The Preliminary Emergency Transfusion Protocol. This is the initial flowchart developed through prior stakeholder interviews asking what an ETP design would look like. This will be iterated to develop a final ETP for clinical use in the hospital.

**Figure 3:**
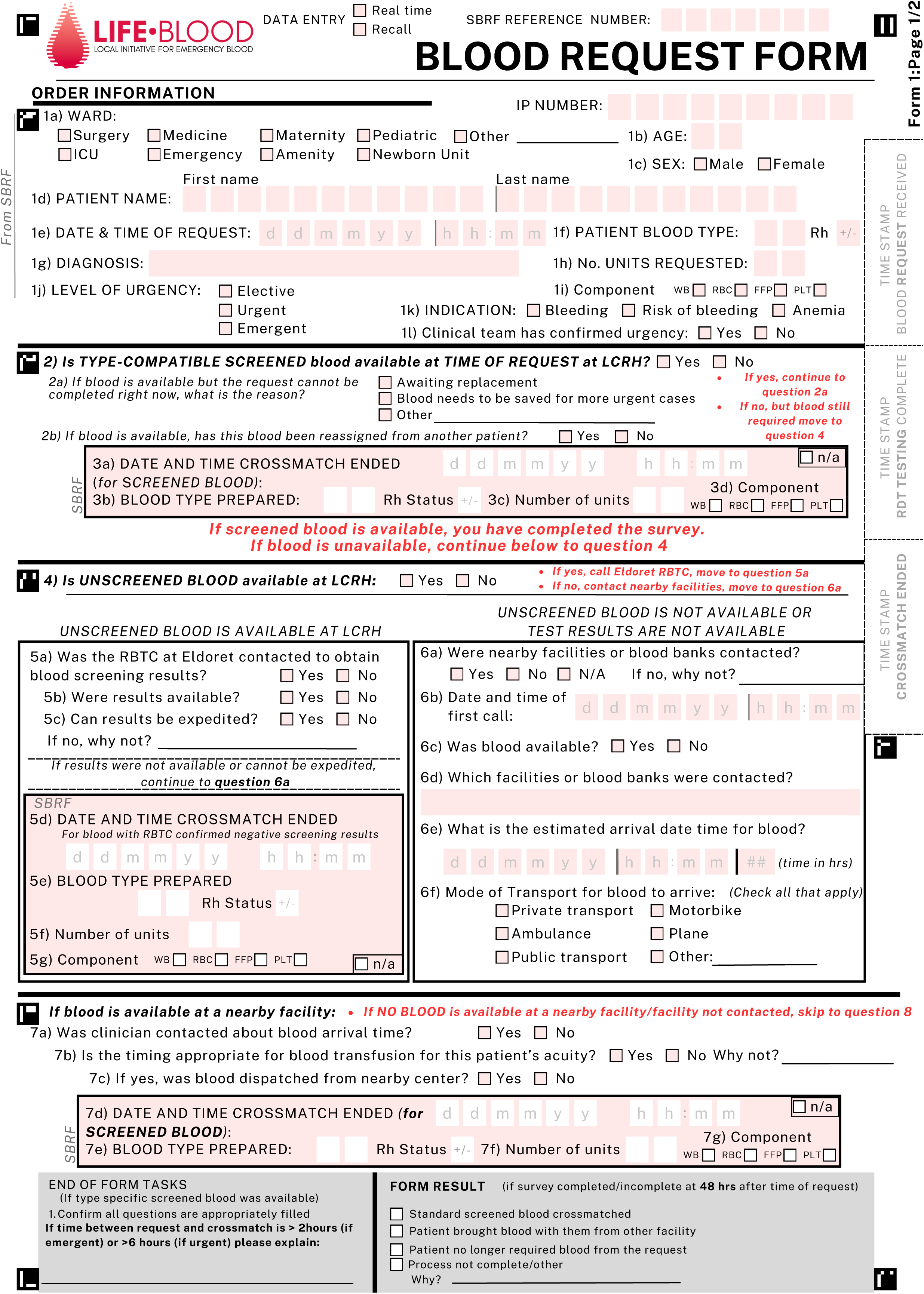
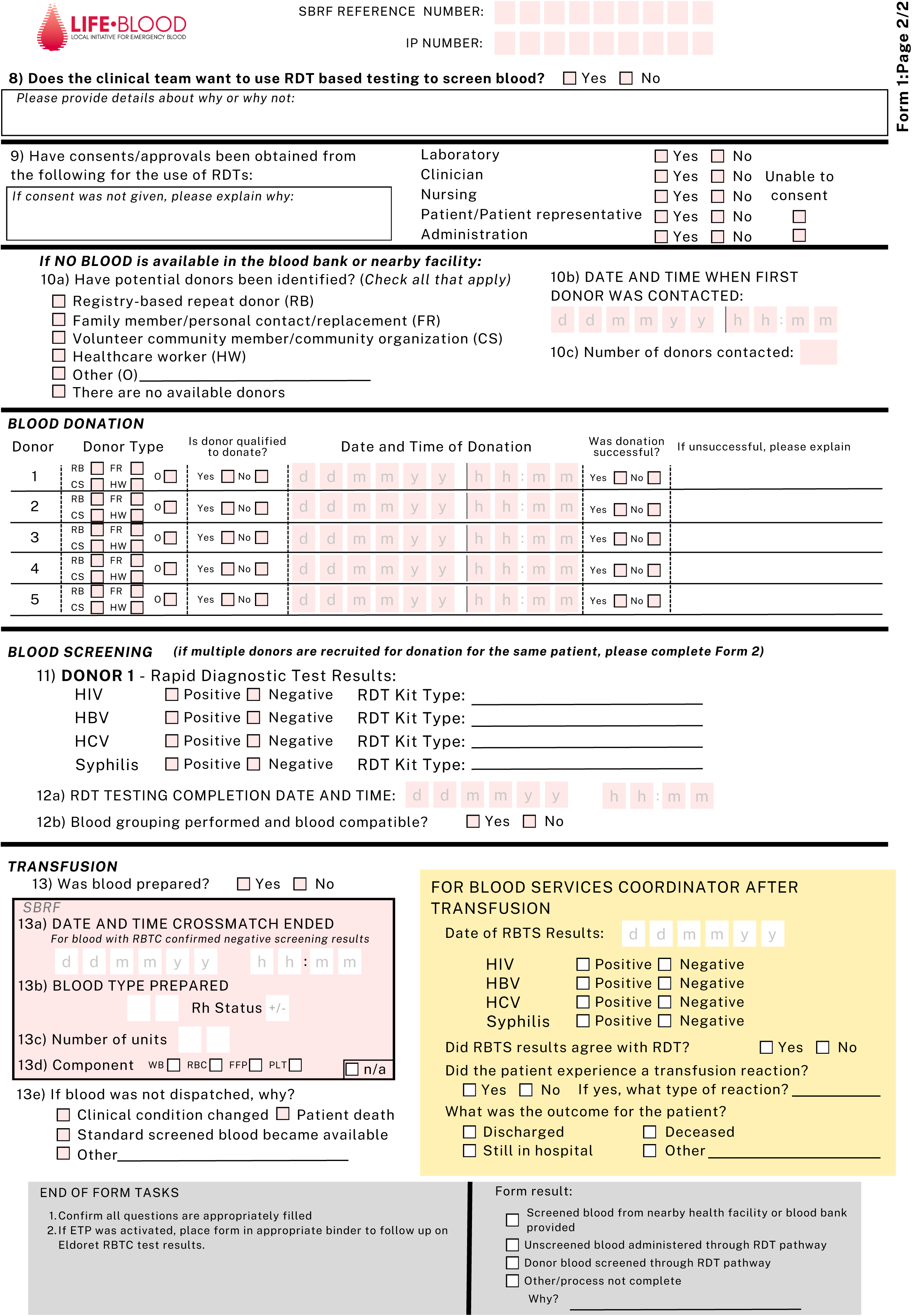
Blood Request Form for ETP Data collection and evaluation of implementation and effectiveness.

We will run a short-term recall survey to provide a daily cross-sectional assessment of blood demand at LCRH. This will occur four times over the study period in one-week increments spanning both pre- and post-ETP initiation. We will send a REDCap based survey via text message to clinical staff caring for patients at LCRH. This survey collects data regarding the number of patients for which blood was requested, how many units were requested, indication, and if there were any patients for which blood was requested in insufficient amounts or not requested at all.

## Data Analysis

The semi-structured interviews will be audio recorded and transcribed using Otter.ai and verified by the research team. In Aim 1, we will use these transcripts as field notes to prepare a finalized ETP for implementation. In Aims 2 and 3, we will analyze transcripts using inductive and deductive qualitative methods to consolidate the data into themes. The research team will first develop general premises into a codebook, and review transcripts iteratively to generate a finalized codebook using MAXQDA Software (2024). Independent reviewers that were not involved in the initial code generation will then code transcripts to thematic saturation with group discussions to resolve disagreements in coding. Final results will present a summary of the themes with representative quotes.

To arrive at consensus regarding CWBB key elements for adaptability, we will use a modified Delphi process. The process will include at least two iterative Delphi rounds to determine the components necessary for a CWBB and components that require local adaptation. We will hold a final round of discussion with the primary study team to finalize the CWBB components. Participants will include multi-level local, regional, and international stakeholders to ensure varied perspectives through regulatory, health system, and government lenses.

For our implementation assessment, we will compile our data into two separate, de-identified databases of 1) retrospective and 2) prospective data. We will determine if there is a change in time to blood dispatch pre- and post-ETP implementation, using a two-sided, two-sample, equal-variance t-test of the prospective data. We will also evaluate clinician perspectives on changes in blood availability and the impact of the ETP.

Based on our prior data, we estimate that there are approximately 20 cases per month of unmet blood requests and of those unmet requests, likely five cases are emergent. We also estimate that the current time to dispatch blood for emergent and unmet blood requests is roughly 12 hours and that this will decrease to six hours after implementation. Given these assumptions, we expect that the emergent, unmet blood request group sample sizes of 12 pre-implementation and 12 post-implementation will achieve 80% power to reject the null hypothesis when the difference in the average time to blood dispatch is 6 hours with a standard deviation of 5 and an alpha of 0.05.

Secondary outcomes will include proportion of urgent (required in less than six hours) unmet blood requests that are met through any means (transport from other facilities, return of regional blood testing results, use of RDT screening); proportion of urgent unmet requests that are met through RDT screening only; number of patients before and after implementation for whom blood was dispatched; units of blood issued, and units of blood collected. In addition, using our retrospective and prospective data we will further characterize changes in time to dispatch, indications for RDT-based transfusions, and proportions of patients with unmet blood needs who were able to receive a transfusion within 24 hours.

## Patient and Public Involvement

To ensure that the results of our study are contextually appropriate and beneficial to the community, we involve local stakeholders at all stages of the research process. We have focused a large portion of our analysis on hospital stakeholder perceptions as this will directly affect their decision-making processes and patient care. To this end, we will directly engage frontline clinicians, nurses, transfusion staff, hospital administrators from LCRH through key informant interviews and group discussions. We will use their feedback to adapt the ETP for the specific needs and operational realities of treating hemorrhage and anemia at LCRH. After the ETP has been employed for several months, hospital stakeholders will be asked about their experience using the ETP and its effect at LCRH on blood availability and decision making around transfusions.

We also wanted to understand Lodwar community perceptions around blood transfusions, blood donations, and the use of an ETP, specifically in emergency settings, as they will be both a major driver for the CWBB as donors and the direct recipient of its care. Previously, patient representatives and blood donors contributed to the initial inquiry of blood transfusion availability. In this part of the study, interviews will explore community-level barriers and facilitators, and cultural and social factors that may influence the willingness to donate or receive blood in the context of the ETP.

Lastly, understanding contextual factors and community needs outside of LCRH will be essential for appropriately adapting WBB to a variety of contexts. Therefore, we will engage local, national, and international health leaders to identify and discuss barriers and facilitators to implementing the ETP. This process will ensure that public input is integral to adapting the protocol for diverse contexts, both within Kenya and internationally.

## Discussion

The global shortage of blood products represents one of the most pressing challenges in healthcare delivery, affecting the management of a broad range of disease states ranging from trauma, obstetrics, malnutrition, infectious disease, oncology, and noncommunicable diseases more broadly. This study represents the first structured implementation and evaluation of a community walking blood bank (CWBB), addressing an urgent need for innovative solutions to chronic blood shortages in remote settings. While various approaches have been proposed to address blood shortages, few have been systematically implemented and evaluated in real-world settings, particularly in blood deserts where the need is most acute.

This study will add important contributions to the understanding of blood transfusion and delivery in resource-limited settings. First, it provides a prospective assessment of actual blood availability - and unavailability - at the local hospital level in a rural context, moving beyond modeled or retrospective analyses that often fail to capture the full scope of the problem or regional analyses that blur the distinction between specific hospital sites. Second, we address the critical issue of hidden demand - where clinicians modify their transfusion practices based on perceived or actual blood availability rather than blood need. By implementing daily provider surveys, we will be the first to quantify this hidden demand, providing crucial insights into the actual scope of blood needs in rural settings. Third, our qualitative analysis examines the impact on healthcare providers, an often-overlooked aspect of blood shortages. Our Phase 1 findings revealed the significant emotional burden on providers, who must choose between using unscreened blood or watching patients die from preventable causes. This study will assess how a structured emergency transfusion protocol affects provider well-being and decision-making. This is particularly relevant given that healthcare worker retention in rural areas is a major challenge for health systems globally, and the stress of managing patients without adequate blood supplies may contribute to burnout and attrition. Fourth, we incorporate patient and family perspectives regarding emergency blood transfusion, providing crucial insights into community acceptance and cultural considerations that may affect implementation.

In order to maximize successful implementation, we have employed the Consolidated Framework for Implementation Research (CFIR) to design the study. This systematic approach accounts for the complex interplay between provider characteristics, patient needs, regulatory requirements, and local resources. The framework helps address several key barriers to CWBB adoption, including implementability, protocol adherence, and maintaining safety standards within an emergency process. Our mixed-methods approach allows us to capture both quantitative metrics of success and qualitative insights into the human factors that will ultimately determine the protocol’s effectiveness.

The regulatory context presents both challenges and opportunities. Currently, the use of RDT for transfusion screening is not permitted in Kenya, as with many other countries. While the existing blood transfusion systems serve most of Kenya effectively, blood desert regions remain (found in almost every country) where the standard system cannot adequately meet transfusion needs. Rather than comparing RDT screening to enzyme immunoassay testing (the current standard of care), we recognize that in true emergencies, the real comparison is between RDT screening and no screening at all - often a choice between life and death. This study aims to formalize a process for providing life-saving blood products while maintaining safety standards when the standard of care is unavailable. Crucially, there is broad support from regulatory authorities and policymakers eager to improve the quality of care available to the populace and reflected in partnerships with the Turkana County Government, Kenya Tissue and Transplant Authority, and the Kenya Ministry of Health. These relationships ensure community support and encourage system improvements through appropriate regulatory channels.

We acknowledge several important limitations of our study. First, this implementation occurs at a relatively unique institution: LCRH has advanced practitioners committed to solving this problem and strong governmental support, an environment that may not be replicated elsewhere. Second, our relatively short data collection window, while powered to assess pre-post implementation changes, may not capture longer-term trends or seasonal variations in blood needs. A multi-site assessment will ultimately be necessary to fully understand the scalability and adaptability of this approach.

The solutions to the blood shortage crisis will require innovative and non-traditional (disruptive) approaches that leverage similarities between contexts while allowing for effective local adaptation. This protocol provides a framework for systematically assessing both the implementation and effectiveness of a CWBB, while gathering crucial data about its potential scalability both within the Turkana region and internationally. By addressing current barriers to adoption and working within existing regulatory frameworks, we hope to provide evidence for a sustainable solution to otherwise preventable deaths due to chronic blood shortages in remote settings.

## Data Availability

There are no data available for this manuscript.

## List of Abbreviations

CWBB: community walking blood bank
EIA: enzyme immunoassay
ETP: emergency transfusion protocol
LCRH: Lodwar County Referral Hospital
LMIC: low- and middle-income countries
RBTC: regional blood transfusion center
TTI: transfusion transmitted infection

## Declarations

### Ethics approval and consent to participate

Ethical approval was granted by the Strathmore University Institutional Scientific and Ethics Review Committee (Approval Reference: SU-ISERC2234/24; Nairobi, Kenya) and the Mass General Brigham (MGB) Hospital’s Institutional Review Board (Protocol #2024P001878, #2024P001879, #2024P001885, and #2024P001887Boston, MA, US). The study team has also secured a research license from the National Commission for Science, Technology, and Innovation (NACOSTI; Reference #168094, Kenya) before initiating the study.

### Availability of data and materials

No datasets have been created for this study. This manuscript does not report data generation or analysis. Future datasets generated during the study will be available from the corresponding author on reasonable request and will follow data management procedures mandated in Kenya by law. The findings will be disseminated through academic publications, conference presentations, and workshops, contributing valuable insights into emergency blood transfusion protocols. Most importantly, these findings will be conveyed to Lodwar County Referral Hospital in order to facilitate quality improvement.

### Author Contributions

NK, NR, TC, and LB conceptualized the design of the study and led submission of the grant application awarded by the Stepping Strong and Laerdal Foundations. FM, RS, AT, CM, AM, SO, EM, NN, KKL, WW, JCP, PK, MD, NR, AC, and TD provided input for the study design, methods, and data analysis procedures. CM, CW, AM, HB, MW, EW, CO, EK, EE, GL, SO, EM, TC provided operational support in obtaining the necessary national, regional, and local approvals to launch the study. NK, FM, RS, AT, NR, TC, and LB drafted the manuscript. All authors critically reviewed, provided feedback on, and approved the final manuscript.

